# Validity and wear compliance of wrist-wearable activity trackers in free-living schoolchildren: validation study

**DOI:** 10.1101/2025.02.20.25322600

**Authors:** Mitsuya Yamakita, Daisuke Ando, Miri Sato, Yuka Akiyama, Kaori Yamaguchi, Zentaro Yamagata

## Abstract

**Introduction:** Wearable activity trackers are widely available for promoting physical activity (PA) and reducing sedentary behavior (SB). However, studies assessing their validity for PA and SB measurement in free-living conditions in school-age children remain limited.

**Objective:** This study aimed to evaluate the criterion validity and wear compliance of the Fitbit Ace, a wrist-worn wearable activity tracker designed for children, in measuring PA and SB under free-living conditions.

**Methods:** A total of 102 children (mean age: 10.2 years; 44.1% girls) simultaneously wore a waist-worn accelerometer (Omron Active Style Pro HJA-750c; ASP HJA-750c) and a Fitbit Ace for 11 consecutive days. Step count and time spent in SB, light PA (LPA), moderate PA (MPA), and vigorous PA (VPA) were collected from both devices. Pearson’s and Spearman’s correlations assessed criterion validity, while Bland–Altman plots, mean absolute percentage error (MAPE), and intraclass correlation coefficients were used to evaluate agreement. Wear compliance was compared between the two devices.

**Results:** The Fitbit Ace showed a very strong correlation with step count and strong correlations with SB, LPA, and moderate-to-vigorous PA (MVPA). However, significant differences were observed for all PA variables. The Fitbit Ace overestimated step count, SB, VPA, and MVPA, whereas it underestimated LPA and MPA. Agreement between devices was low, with proportional errors in step count, LPA, VPA, and MVPA. MAPE exceeded 20% for all variables, including step count (21.7%), SB (31.2%), LPA (30.3%), and MVPA (120.7%). The proportion of children meeting the criterion of ≥10 h/day wear criterion was higher for the Fitbit Ace (83.2%) than for the ASP HJA-750c (57.8%), as was adherence to ≥4 days (97.0% vs. 62.2%).

**Conclusion:** While caution is needed when using the Fitbit Ace for precise measurement of individual PA, it is a useful tool for assessing overall patterns of step count, SB, and LPA in school-aged children.

## Introduction

Physical activity (PA) provides numerous health-related benefits [1]. It is known to enhance cognitive skills and prosocial behavior among children and adolescents [2]. Recent evidence shows that many health and social benefits extend into adulthood [1,3]. Additionally, there is emerging evidence of the negative health effects and potential public health burden associated with excessive sedentary behavior (SB) [4]. However, 81% of adolescents aged 11–17 years are insufficiently physically active, and excessive sedentary time is also widespread among children and adolescents globally [5, 6]. Thus, urgent bolstering of effective policies and programs to increase population PA levels, including in children and adolescents, is needed [1,7].

Among the various behavior change techniques used to promote PA, self-monitoring of health behavior is reported to be effective and highly appreciated in changing PA levels [8,9]. These findings suggest that PA and SB surveillance using accurate evaluation tools and PA promotion via cost-effective interventions in school-aged children are top public health priorities [10]. Despite the general consensus that research-based accelerometers are the gold standard for measuring habitual PA in surveillance and epidemiologic research [11], the use of accelerometers entails relatively high costs, has low wearing compliance, and extensive training for data processing, and does not lead to the maintenance of long-term monitoring after the survey and study is completed [12,13].

With these concerns, consumer-based PA monitors, often referred to as “wearable activity trackers,” are widely available on the market and recent scientific evidence supports wearable activity trackers might increase daily steps in children and adolescents [14,15]. Therefore, wearable activity trackers have great potential for substituting research-based accelerometers in clinical and epidemiologic research [14,15]. In particular, wrist-worn wearable activity trackers have been shown to offer better compliance when compared with waist-worn devices because they are easy to wear, comfortable, and waterproof, reducing participant burden. Therefore, users may prefer them and wear them continuously day and night [14–16]. Fitbit (Fitbit Inc., San Francisco, CA, USA) is one of the most popular commercial wearable activity trackers [17], and many studies have shown that the use of Fitbit devices in interventions may promote PA [18]. A systematic review assessed the accuracy of 72 wrist-wearable activity trackers and reported consistent accuracy in step counts for the Fitbit Charge and Fitbit Charge HR [19]. However, studies evaluating PA and SB in children using the Fitbit Ace, developed to assess activity in children aged 8 years and older in a free-living environment, are lacking.

To our knowledge, the Fitbit Ace has only been evaluated in studies that assessed step counts under experimental conditions (one of which used the Fitbit Ace 2, the successor to the Fitbit Ace) [20, 21], and one study that evaluated the validity of Fitbit Ace 2–derived step counts and moderate-to-vigorous physical activity (MVPA) under free-living conditions [22]. Neither of these studies adequately demonstrated the validity of estimating MVPA and step count. Moreover, no studies have investigated SB or light physical activity (LPA) or examined the difference in wear compliance when compared with a waist-worn accelerometer. The widespread use of consumer-wearable activity trackers for children could help develop effective interventions to promote PA and reduce SB levels among school-aged children [15, 23]. Therefore, in this study, we aimed to evaluate the criterion validity, including SB and LPA, and wear compliance of the Fitbit Ace, for PA assessment under free-living conditions in school-aged children.

## Materials and methods

### Study design

This validation study used a baseline survey from the Koshu GRoup Activity, Play, and Exercise (GRAPE) study. The Koshu GRAPE study was a school-based cluster randomized controlled trial that examined whether short-time exercise increases bone stiffness, PA, SB, and non-cognitive skills conducted from 2018 to 2019 (Trial registration number: UMIN-CTR UMIN000034992) [24].

### Participants

Participants were recruited from two of the nine schools that participated in the Koshu GRAPE study. This study included 107 children in the 4th and 5th grades (aged 9–11 years) from the 2018 school year.

This validation study was approved by the ethics committee of the Faculty of Medicine, University of Yamanashi (approval no. 1929) and was conducted with the cooperation of the Health Promotion Division and the Board of Education of the Koshu City Administration Office. The principals of the participating schools provided ethical approval. All participants provided informed assent to participate, and written informed consent was obtained from their guardians.

### Sample size

The sample size was 20 participants calculated with power (β=0.8), error (α=0.05), and correlation (r=0.6, strong [25]). Furthermore, we assumed that at least 64 children were needed, based on our previous studies that targeted children in Koshu City [24], considering the accept rate (90%), missing due to invalid accelerometers (30% invalid [26]), and considering sex-stratified analysis for wear compliance assessment. Since no school had more than 64 children in the 4th and 5th grades alone, two large schools that exceeded the criteria (107 children in total) were included in the study. Instruments

### Omron Active style Pro HJA-750c

The Omron Active Style Pro HJA-750c (ASP HJA-750c; Omron Healthcare, Kyoto, Japan) is a waist-worn tri-axial accelerometer with dimensions of 52 × 40 × 12 mm and a weight of approximately 23 g, including the battery. It was used as the reference device for measuring PA and SB in this study. The Active style Pro 350 IT, the same acceleration sensor and algorithm as the 750c [27–29], has been validated as well as or better than accelerometers widely used in Western countries for measuring PA and SB [30,31]. The ASP HJA-750C has also been validated for PA measurement in adults [29] and step count in children [32] and is used in daily activity measurement in children of a wide age range [33–35].

### Fitbit Ace

The Fitbit Ace is a wrist-worn activity tracker designed to monitor PA and sleep in children aged 8 years and older. It features a triaxial accelerometer and continually acquires data with an onboard storage capacity for approximately five days without syncing. Data were transferred via Bluetooth to the Fitbit application program interface (API) using Fitbit’s mobile app.

### Procedure

The participants were instructed to wear the ASP HJA-750c and Fitbit Ace simultaneously for 11 days (four weekend days [two weekends] and seven weekdays [five days in between two weekends and one day before and after two weekends]). The participants were asked to wear the ASP HJA-750c on the right side of the waist during waking time, except during water-based activities (bathing, swimming, etc.) and while playing activities with a high risk of injury, and to wear the Fitbit Ace on the nondominant wrist throughout the day in principle, including sleep, unless it was in the way or posed a risk of injury or other danger when worn.

The participants’ age, height, and weight were collected via physical measurements taken during medical checkups conducted at elementary schools in January 2019. Body mass index (kg/m^2^) was calculated using height and weight, and obesity was categorized based on the study software of the Joint Committee on Standard Values of the Japan Pediatric Endocrine Society and the Japan Society for Growth Research [36].

## Data processing

### ASP HJA–750c data processing

The ASP HJA–750c CSV data files were downloaded using the Omron health management software BI-LINK for PA professional edition ver1.0. When the ASP HJA– 750c was used to evaluate PA and SB in primary school children, metabolic equivalents (METs) values were overestimated [37] because the predictive equations were established for adults. Therefore, we created a Python script to calculate the METs for children using the recommended conversion equations for primary school students [37,38] and converted the CSV data for children. A macro program (ver. 1.1), developed and distributed by the Japan Physical Activity Research Platform [38], was used for CSV data processing. Data were collected in 1-min epochs. Activity behaviors were classified into four intensity categories based on metabolic equivalents (METs): SB (≤1.5 METs), LPA (1.6–2.9 METs), moderate PA (MPA, 3.0–6.0 METs), and vigorous PA (VPA, ≥6.0 METs) [39].

### Fitbit Ace data processing

Fitbit accounts of the participants were created and set up in advance by the researcher, and pre-configured devices were distributed to the children. Data from the Fitbit Ace are automatically stored on the Fitbit server when synchronized via Bluetooth. However, because the children themselves were unable to use the Fitbit App at school and the built-in device can only store 5 days of measurement data, the researchers visited the school once every 3 days during the 11-day measurement period and used their own smartphones and tablets with the Fitbit app to synchronize and obtain the data. We then requested TechDoctor Inc. to retrieve and download the data using the Fitbit API. A Python script was used to collect the data in 1-min intervals and aggregate the data. “Sedentary,” “Light active,” “Moderately/Fairly active,” and “Very active” activity intensity levels from the Fitbit were considered as sedentary (≤1.5 METs), light (1.5–3.0 METs), moderate (3.0–6.0 METs), and vigorous (≥6.0 METs) activity, respectively [40].

### Extraction of valid data and outcomes

For both devices, ≥60 consecutive min with accelerometer non-response was defined as non-wear time, and the wear time was calculated by subtracting the non-wear time from 24 h. However, since the Fitbit Ace was requested to be worn during sleep, the wearing time was defined as the time from waking to bedtime if sleep time was recorded, or if not recorded, as the waking time on weekdays and weekends at 6:30 and 7:00, respectively, and the bedtime on weekdays and the day before weekends at 21:30 and 22:00, respectively, based on a study of children in Koshu City [41].

### Validation assessment

We defined valid person-days as if both devices had valid days (≥10 h of wear) and the difference in wear time between the devices was ≤60 min [42]. We evaluated the steps, time spent in each PA intensity (LPA, MPA, VPA, and MVPA), and SB on person-days.

### Wear compliance assessment

The outcomes of interest were the percentage of children as ≥10 h of wear time per day and ≥4 valid days during the 11-day measurement period (at least three weekdays and one weekend) [43,44]. As other valid wear time criteria, we also examined the percentage of person–days meeting ≥8 h, 12 h, and 14 h of wear time per day and the percentage of children meeting two days regardless of weekdays [45] and weekends, ≥3 valid days (at least two weekdays and one weekend [46]), ≥5 valid days (at least four weekdays and one weekend, and at least three weekdays and two weekends), ≥6 valid days (at least five weekdays and one weekend, and at least four weekdays and two weekends), and ≥7 valid days (at least six weekdays and one weekend, and at least five weekdays and two weekends).

## Statistical analysis

### Validity assessment

Criterion validity was determined using Pearson’s correlations between step counts, PA, and SB estimates from the ASP HJA-750c and Fitbit Ace. Because MPA, VPA, and MVPA had skewed distributions with a long right tail, Spearman’s rank sum test was adopted. The following cut-off values were used to interpret the Pearson’s and Spearman’s correlation: r<0.20= negligible, 0.20–0.39=weak, 0.40–0.59=moderate, 0.60– 0.79=strong, and 0.80–1.0=very strong.

Agreement was assessed using the intraclass correlation coefficient (ICC) analysis (2-way random, absolute agreement). [47]. The cut-off values used to interpret the ICC were <0.20=poor, 0.21–0.40=Fair, 0.41–0.60=moderate, 0.61–0.80=substantial, and 0.81–1.00=almost perfect [47]. Bland–Altman plots were used to visualize the agreement between the ASP HJA-750c and Fitbit Ace. Proportional bias was assessed by examining the linear association between the measurements of the two devices. Additionally, Student’s t-test was used to compare the differences in measurements between the two devices, and the mean absolute percentage error (MAPE) was calculated to evaluate the similarity of the estimates. MAPEs were calculated by taking the absolute difference between the ASP HJA-750c values and the Fitbit Ace value, dividing it by the ASP HJA-750c value, expressing the result as a percentage, and then averaging it across all data points. The MAPE was categorized based on commonly used accuracy cutoffs for measuring the step count with wearable devices, with an acceptable error of ±10% in free-living settings [48]. Our categorization for the MAPE was therefore as follows: poor (>20%), moderate (10.1–20%), good (3.1–10%), and excellent (0–3%).

### Wear compliance assessment

Student’s t-test was used to compare the means of wearing time throughout the measurement period between the ASP HJA-750c and Fitbit Ace. The percentages that met the wear time per person–day (8 h, 10 h, 12 h, and 14 h) and the percentages that met the reasonable number of days required to estimate habitual PA (2–7 days) were compared using a Chi-square test.

All statistical analyses were performed using STATA 18 SE (Light Stone), and an alpha level of 0.05 was set to define statistical significance for all analyses.

## Results

### Participants and valid measurement person-days

A total of 102 children agreed to the study (consent rate: 95.3%). Participants’ characteristics are presented in Table 1. The proportion of girls was 44.1%, and the average age of the children was 10.2 years (SD = 0.7).

**Table 1.** Participants Characteristics by Sex Among Japanese School-Aged Children.

|  | **Total** |  | **Boys** |  | **Girls** |  |
| --- | --- | --- | --- | --- | --- | --- |
|  | **n = 102** |  | **n = 57** |  | **n = 45** |  |
| **Age, years (SD)** | 10.2 | (0.7) | 10.1 | (0.7) | 10.2 | (0.6) |
| **Height, cm (SD)** | 140.1 | (7.2) | 139.2 | (7.2) | 141.3 | (7.3) |
| **Weight, kg (SD)** | 35.5 | (7.7) | 35.6 | (8.7) | 35.3 | (6.4) |
| **BMI, kg/m<sup>2</sup> (SD)** | 17.9 | (2.6) | 18.2 | (3.0) | 17.6 | (2.1) |
| **Weight status, n (%)** |  |  |  |  |  |  |
| Underweight | 2 | (2.0) | 1 | (1.8) | 1 | (2.2) |
| Healthy weight | 84 | (82.3) | 44 | (77.2) | 40 | (88.9) |
| Overweight | 12 | (11.8) | 8 | (14.0) | 4 | (8.9) |
| Obese | 4 | (3.9) | 4 | (7.0) | 0 | (0.0) |

A total of 1,122 person-days of measurements over 11 days for 102 participants were included, and 135 person-days (12%) met the criteria for validity assessment (wearing at least 10 h per day, with a difference of ≤60 min for the two devices).

For ASP HJA-750c, 1,053 person-days (93.9%) were included in the analysis, excluding four children who had not taken all 11 days (44 person-days) and 11 children with device water immersion, device malfunction, device loss, or device battery trouble (25 person-days). For Fitbit Ace, 83 children could not be measured on the first day (83 person-days); three (9 person-days) due to out-of-battery or device malfunction, and two where data could not be obtained from the API for unknown reasons (22 person-days) were excluded; finally, 1,008 person-days (89.8%) were included in the analysis.

### Criterion validity

A very strong correlation between the two devices was found for step count (r=0.856, p<0.001). Strong correlations were also found for SB and LPA. However, the correlation was moderate for MPA, VPA, and MVPA (Table 2).

**Table 2.**
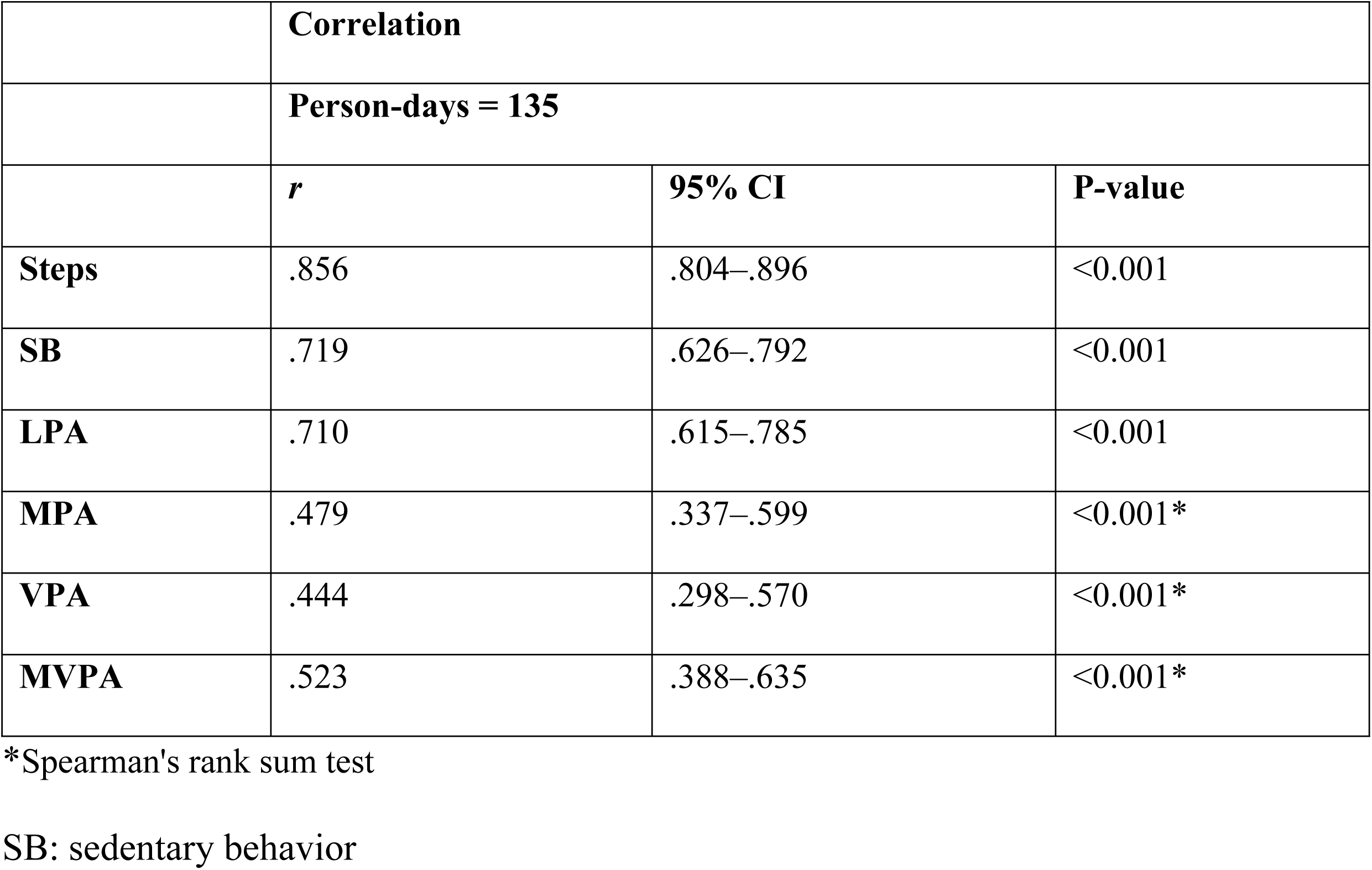
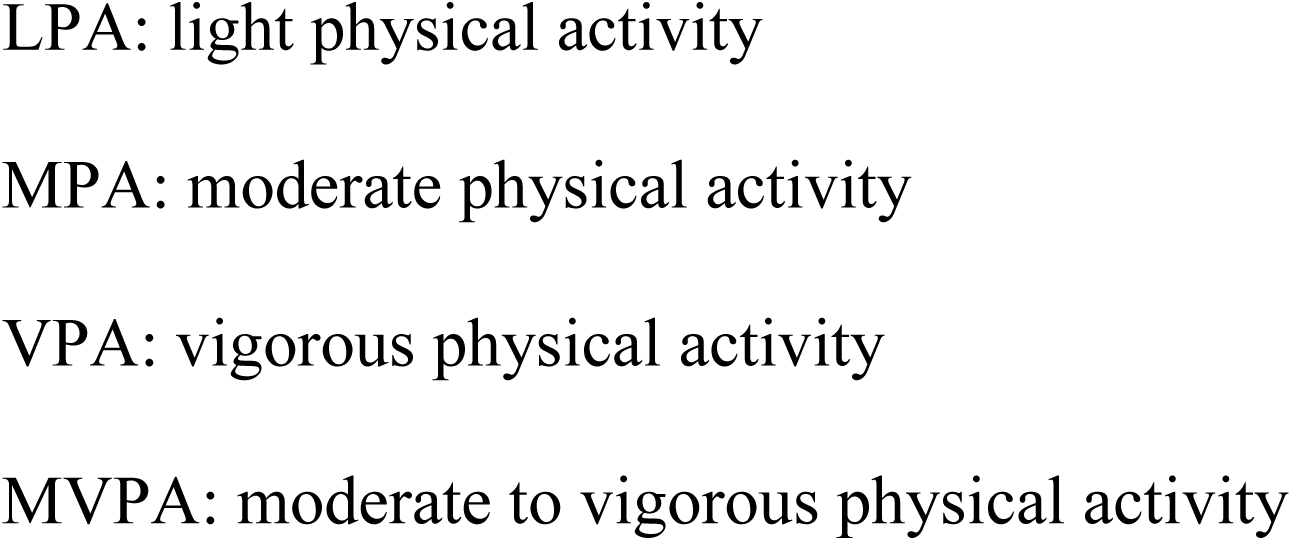
Criterion Validity Based on Pearson’s and Spearman’s Correlation Coefficients Between the ASP HJA-750c and Fitbit Ace.

|  | Correlation |  |  |
| --- | --- | --- | --- |
|  | Person-days = 135 |  |  |
|  | *r* | 95% CI | P-value |
| **Steps** | .856 | .804–.896 | <0.001 |
| **SB** | .719 | .626–.792 | <0.001 |
| **LPA** | .710 | .615–.785 | <0.001 |
| **MPA** | .479 | .337–.599 | <0.001* |
| **VPA** | .444 | .298–.570 | <0.001* |
| **MVPA** | .523 | .388–.635 | <0.001* |

\*Spearman's rank sum test
SB: sedentary behavior

Table 3 shows the differences between the values obtained using the two devices. Significant differences were found between the two devices for all PA variables. The Fitbit Ace recorded higher values for step count, SB, MPA, and MVPA, whereas lower values were recorded for LPA and MPA. Notably, a significant difference was observed in the recording of VPA between the two devices. The MAPE exceeded 20% for all the variables: the smallest value was found for step count (approximately 21%), whereas the highest value was found for MVPA (120%). For VPA, the MAPE could not be calculated because of the inclusion of zeros.

**Table 3.**
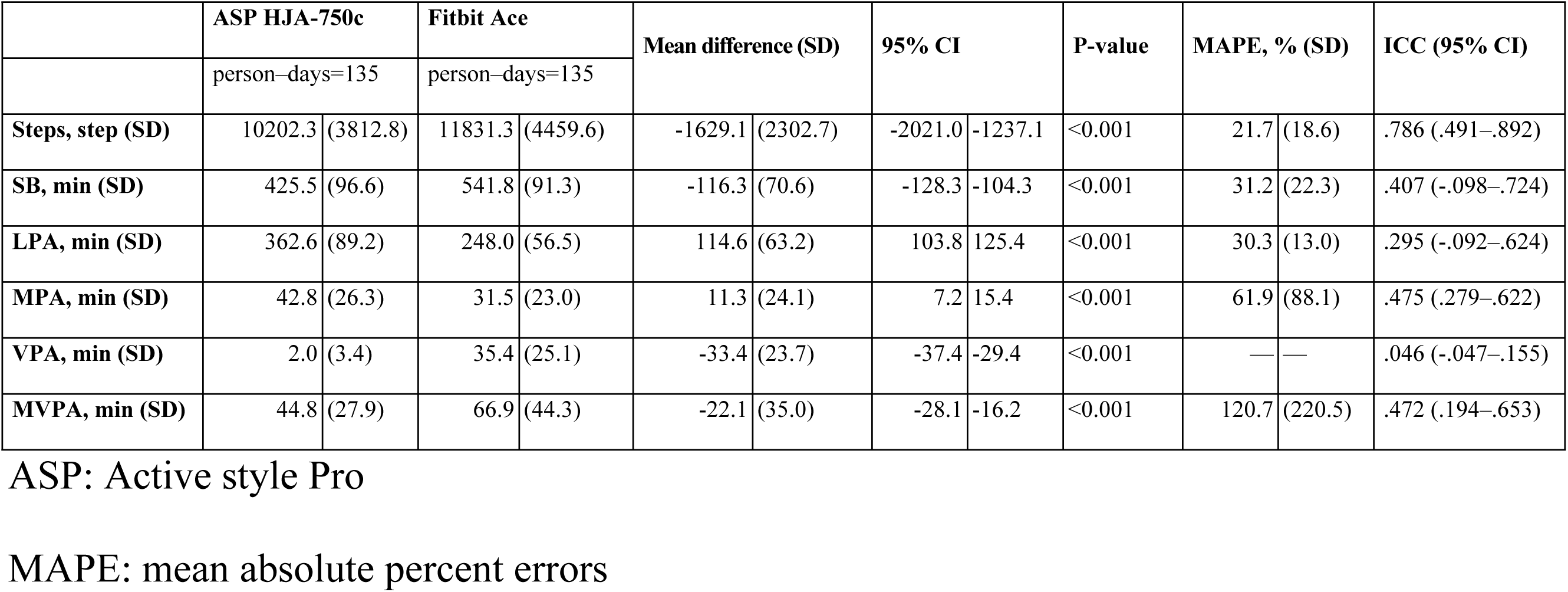
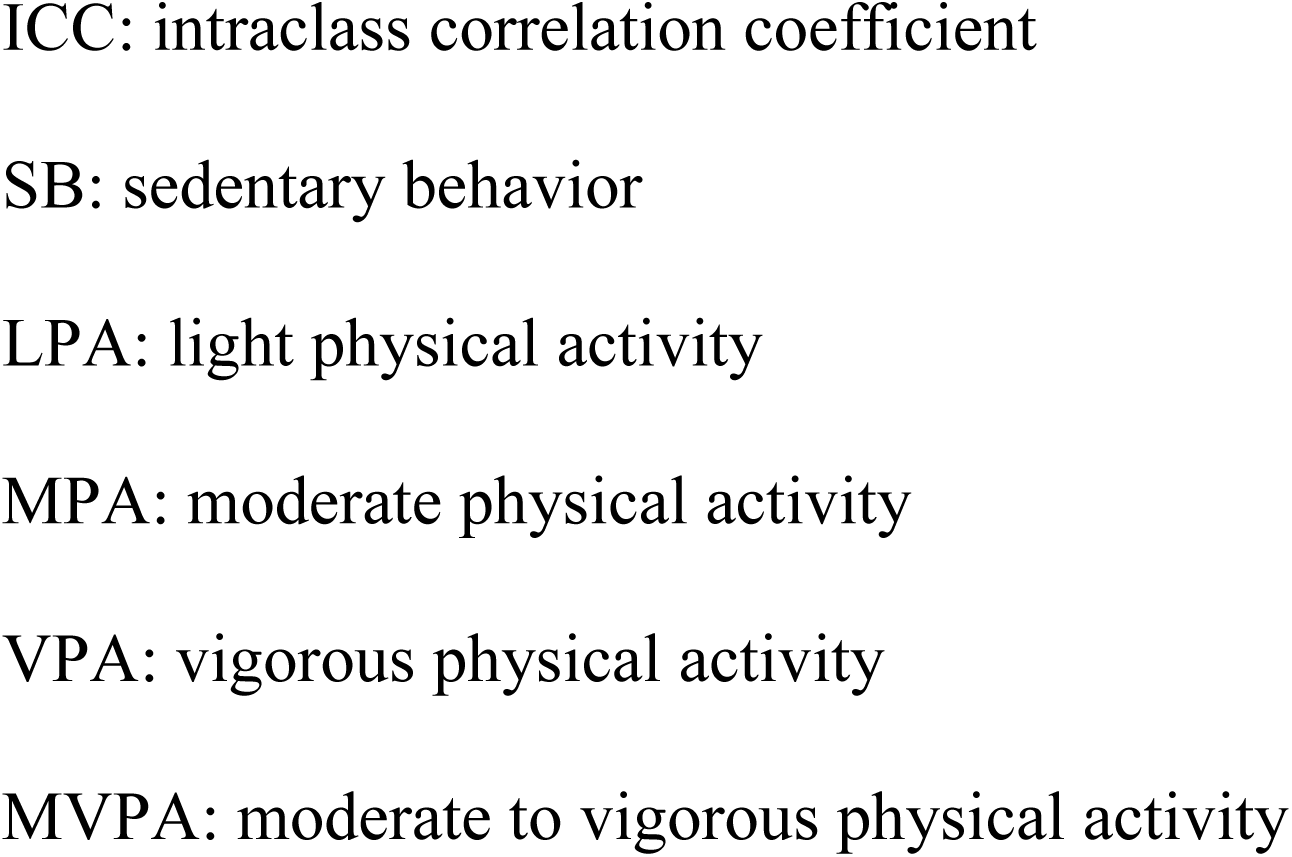
Mean Absolute Percent Errors and Intraclass Correlation Coefficient of Steps, SB, and Each PA Intensity Between the ASP HJA-750c and Fitbit Ace.

Fig 1 shows the agreement between the two devices using a Bland–Altman plot. The Fitbit Ace overestimated the step count, SB, VPA, and MVPA compared to the ASP HJA-750c, whereas it underestimated LPA and MPA. Substantial discrepancies were observed in SB, LPA, and VPA measurements. The majority of plots for SB and VPA fell below zero, which indicates that the Fitbit Ace systematically overestimated these variables compared to the ASP-750c. Conversely, most LPA plots were positioned above zero, which shows that the Fitbit Ace consistently underestimated LPA relative to the ASP-750c.

**Fig 1.**
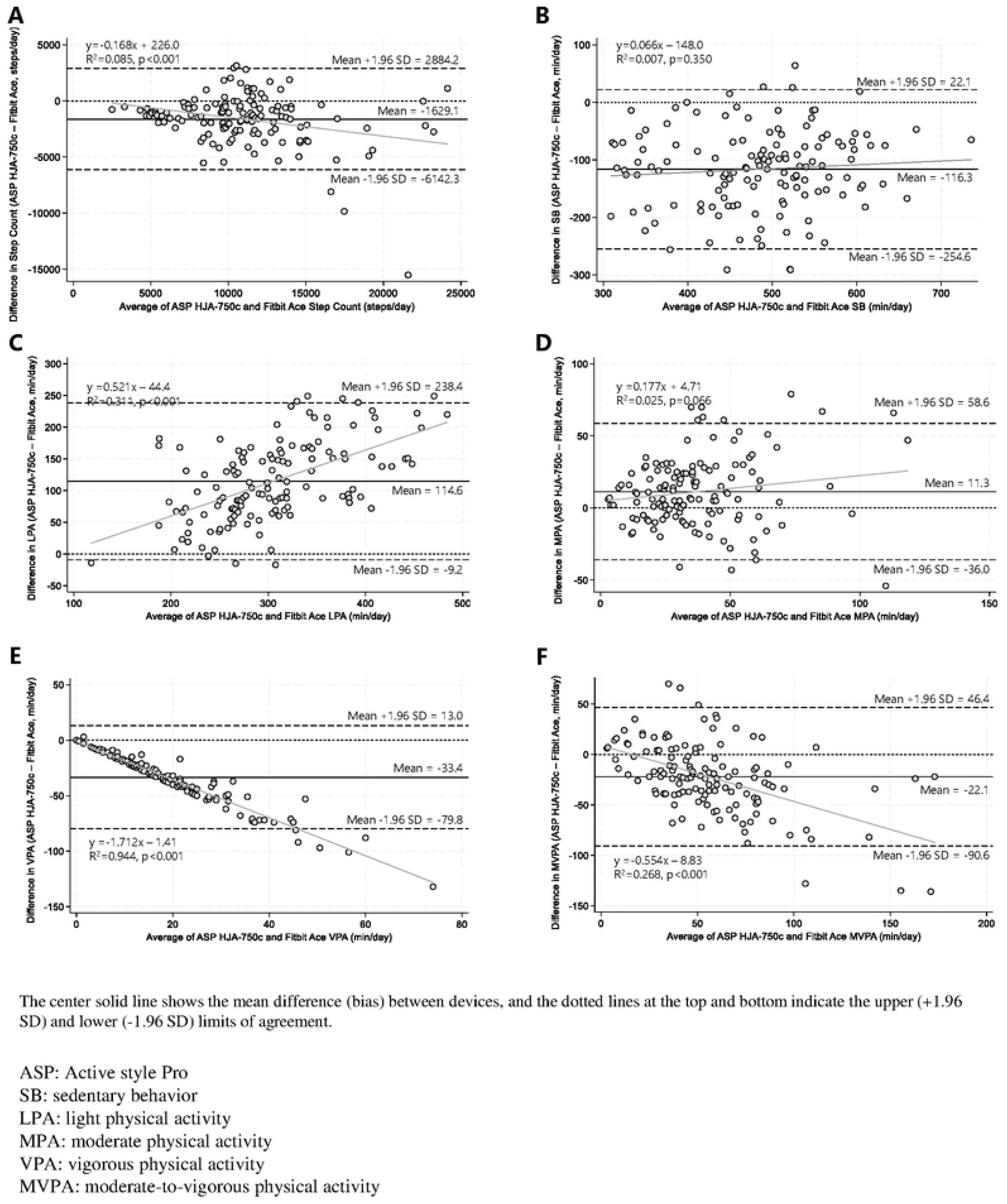
Bland–Altman Plots Visualizing Agreement Between ASP HJA–750c and Fitbit Ace–Derived Step Counts (A), SB (B), LPA (C), MPA (D), VPA (E), MVPA (F). The solid line in the center shows the mean difference (bias) between the devices, and the dotted lines at the top and bottom indicate the upper (+1.96 SD) and lower (–1.96 SD) limits of agreement. ASP: Active style Pro; SB: sedentary behavior; LPA: light physical activity; MPA: moderate physical activity; VPA: vigorous physical activity; MVPA: moderate-to-vigorous physical activity

Furthermore, the results of the regression analysis showed that proportional errors were observed in the step count, LPA, VPA, and MVPA, where the larger the value, the greater the discrepancy (in the negative direction for LPA and in the positive direction for VPA and MVPA).

### Wear compliance

The average daily wearing time was 9.4 h/person–day for the ASP HJA-750c and 11.9 h/day for the Fitbit Ace, with the Fitbit Ace being worn significantly longer overall and stratified by sex (Table 4). Using 8 h, 10 h, 12 h, and 14 h of wear time criteria person–days, the Fitbit Ace compliance was higher than the ASP HJA-750c compliance for all wear time criteria. This association remained the same during the examination of boys and girls.

**Table 4.** Comparison of Wear Compliance Between the ASP HJA-750c and Fitbit Ace.

|  | Total |  |  |  |  | Boys |  |  |  |  | Girls |  |  |  |  |
| --- | --- | --- | --- | --- | --- | --- | --- | --- | --- | --- | --- | --- | --- | --- | --- |
|  | ASP HJA-750c |  | Fitbit Ace |  | P-value | ASP HJA-750c |  | Fitbit Ace |  | P-value | ASP HJA-750c |  | Fitbit Ace |  | P-value |
|  | Person-days<br>= 1,053 |  | Person-days<br>= 1,008 |  |  | Person-days<br>= 581 |  | Person-days<br>= 561 |  |  | Person-days<br>= 472 |  | Person-days<br>= 447 |  |  |
| Total wear time,<br>min/person-days (SD) | 9.4 | (5.9) | 11.9 | (4.7) | <0.001 | 9.4 | (6.0) | 11.6 | (4.9) | <0.001 | 9.3 | (5.8) | 12.3 | (4.5) | <0.001 |
| Valid wear hours,<br>person-days (%) |  |  |  |  |  |  |  |  |  |  |  |  |  |  |  |
| ≥8 h | 693 | (65.8) | 902 | (89.5) | <0.001 | 377 | (64.9) | 492 | (87.7) | <0.001 | 316 | (67.0) | 410 | (91.7) | <0.001 |
| ≥10 h | 609 | (57.8) | 839 | (83.2) | <0.001 | 330 | (56.8) | 458 | (81.6) | <0.001 | 279 | (59.1) | 381 | (85.2) | <0.001 |
| ≥12 h | 463 | (44.0) | 738 | (73.2) | <0.001 | 258 | (44.4) | 398 | (70.9) | <0.001 | 205 | (43.4) | 340 | (76.1) | <0.001 |
| ≥14 h | 232 | (22.3) | 515 | (51.1) | <0.001 | 130 | (22.4) | 269 | (48.0) | <0.001 | 102 | (21.6) | 246 | (55.0) | <0.001 |

ACTIVITY TRACKER VALIDITY AND WEAR COMPLIANCE IN CHILDREN
|  | n = 98 |  | n = 100 |  |  | n = 54 |  | n = 56 |  |  | n = 44 |  | n = 44 |  |  |
| --- | --- | --- | --- | --- | --- | --- | --- | --- | --- | --- | --- | --- | --- | --- | --- |
| Valid wear days, n (%) |  |  |  |  |  |  |  |  |  |  |  |  |  |  |  |
| ≥2 days | 90 | (91.8) | 99 | (99.0) | 0.016 | 48 | (88.9) | 56 | (100.0) | 0.010 | 42 | (95.5) | 43 | (97.7) | 0.557 |
| ≥3 days (2 WD and 1 WE) | 64 | (65.3) | 98 | (98.0) | <0.001 | 35 | (64.8) | 55 | (98.2) | <0.001 | 29 | (65.9) | 43 | (97.7) | <0.001 |
| ≥4 days (3 WD and 1 WE) | 61 | (62.2) | 97 | (97.0) | <0.001 | 33 | (61.1) | 55 | (98.2) | <0.001 | 28 | (63.6) | 42 | (95.5) | <0.001 |
| ≥5 days (4 WD and 1 WE) | 58 | (59.2) | 94 | (94.0) | <0.001 | 31 | (54.4) | 53 | (93.0) | <0.001 | 27 | (60.0) | 41 | (91.1) | 0.001 |
| ≥5 days (3 WD and 2 WE) | 45 | (45.9) | 89 | (89.0) | <0.001 | 23 | (40.4) | 49 | (86.0) | <0.001 | 22 | (48.9) | 40 | (88.9) | <0.001 |
| ≥6 days (5 WD and 1 WE) | 48 | (49.0) | 85 | (85.0) | <0.001 | 26 | (45.6) | 47 | (82.5) | <0.001 | 22 | (48.9) | 38 | (84.4) | <0.001 |
| ≥6 days (4 WD and 2 WE) | 43 | (43.9) | 87 | (87.0) | <0.001 | 22 | (38.6) | 47 | (82.5) | <0.001 | 21 | (46.7) | 40 | (88.9) | <0.001 |
| ≥7 days (6 WD and 1 WE) | 35 | (35.7) | 72 | (72.0) | <0.001 | 18 | (31.6) | 36 | (63.2) | 0.001 | 17 | (37.8) | 36 | (80.0) | <0.001 |
| ≥7 days (5 WD and 2 WE) | 36 | (36.7) | 79 | (79.0) | <0.001 | 19 | (33.3) | 42 | (73.7) | <0.001 | 17 | (37.8) | 37 | (82.2) | <0.001 |

ASP: Active style Pro
WD: weekdays
WE: weekend days

For the percentage of days meeting the criteria of ≥4 valid days, Fitbit Ace was significantly greater than ASP HJA-750c, with 97% meeting that criterion. Although the difference between the two devices disappeared for the criterion of ≥2 valid days for girls, the Fitbit Ace met the criteria more frequently than the ASP HJA-750c in the other criteria (Table 4).

## Discussion

To the best of our knowledge, this is the first study in which the validity and wear compliance of the Fitbit Ace has been simultaneously examined in school-aged children in free-living conditions. We found that the Fitbit Ace measurements were strongly correlated with step count, SB, and LPA and moderately correlated with MPA, VPA, and MVPA.

However, the agreement between the two devices was low, and the MAPE compared to the ASP-750c exceeded 20% for all indicators, especially for VPA (120%). Additionally, the Fitbit Ace was found to overestimate the step count, VPA, and MVPA and underestimate SB and LPA compared to the ASP-750c. Proportional errors were identified for step count, LPA, VPA, and MVPA, with larger errors for larger values. However, extremely high wear compliance was observed for all indices compared with the ASP-750c waist-worn accelerometer.

These findings suggest that while the Fitbit Ace should be used with caution for precise individual PA measurement, its strong correlation with step count and LPA, combined with excellent wear compliance, support its effectiveness as a tool for monitoring overall trends in children’s step counts and light-intensity PA patterns.

### Validity assessment

The step count recorded by the Fitbit Ace was highly correlated with and overestimated by the waist-worn accelerometer. This finding is consistent with many previous studies that consumer model trackers such as Fitbit are the most accurate in measuring step count but have insufficient validity for measuring MVPA [49]. However, our results contradict those of a study that used the same Fitbit Ace, in which step count was underestimated [20]. This discrepancy may be due to the use of direct step counting as the gold standard method and the evaluation being conducted under experimental conditions. The step count measured by accelerometers has been shown to be underestimated when compared with direct observations [50].

The results for MPA, VPA, and MVPA revealed moderate correlations but low agreement. These findings are consistent with previous studies on wrist-worn wearable devices [49] and the successor of the Fitbit Ace, the Fitbit Ace 2 [21]. In particular, although the MAPE of VPA indicated considerable disagreement, the result aligns with a previous study that compared data from the Fitbit Flex 2—an activity tracker with functionality similar to the Fitbit Ace—with measurements obtained from the waist-worn ActiGraph GT9X accelerometer [51]. In contrast, the results for LPA differed from those of our study, showing low correlation and agreement [51]. The different validity criteria used in these studies (e.g., wearing time >8 h) may have contributed to these discrepancies.

For SB, Byun et al. [52] reported a high correlation (r = 0.87, Pate’s criterion; r = 0.85, Everson’s criterion), similar to our findings. By contrast, Schmidt et al. [51] reported moderate correlations. These discrepancies may also be attributable to differences in the criteria for wearing time.

In our analysis, the difference in wear time between the two devices had to be within 60 minutes. However, Byun et al. aligned their data in 1-minute increments, ensuring no difference in wear time [52]. In comparison, Schmidt et al. included data when both devices were worn for at least 8 hours per day [51], which may have contributed to the lower correlation. Although Fitbit activity trackers (such as the Fitbit Zip) may overestimate SB, evidence suggests they are valid tools for measuring SB among children and young populations [53]. The Fitbit Ace may exhibit similar measurement properties.

### Wear compliance

Herein, the widely used validity criterion of at least 10 h wear time per day for 4 days (including one weekend day) was met in 97% of cases for the Fitbit Ace compared to 62% for the ASP HJA-750c, which indicates that the Fitbit Ace had higher wear compliance than the waist-mounted accelerometer did. This finding is consistent with those of previous studies conducted among children on similar wrist-worn Fitbit devices [54,55] and other valid wrist-worn accelerometers (ActiGraph [56], GENEActive [57,58]) that support higher wear compliance with wrist-worn devices. In addition, approximately 80% of the children met the requirement of wearing the Fitbit Ace for at least 7 days (five weekdays and two weekend days, representing typical physical activity for a week). This suggests that the Fitbit Ace is a suitable device for long-term measurement of daily PA among school-aged children.

However, the percentage of valid data based on daily wear time decreased to 73% when the validity criterion was set at ≥12 h and to nearly 50% when the criterion was set at ≥14 h. Wing et al. [55] also reported that proportion of valid data decreased as the criteria for daily wear-time increased (73% at 10 h, 58% at 12.5 h, and 21% at 15 h) which is consistent with our findings. Given that high compliance allows for the measurement of validated daily activity over a shorter period and enhances confidence in the representativeness of daily PA [59], our findings suggest that the Fitbit Ace is useful for long-term activity measurement in school-aged children.

### Strengths and limitations

The strengths of this study include that it is the first study to investigate simultaneously the validity of all PA intensities, including SB and LPA, and the wear compliance of the Fitbit Ace under free-living conditions over an 11-day measurement period, including two weekends, with a sufficient sample of children. Our study provides novel findings that address vital gaps in scientific literature and underscores the utility of wrist-worn wearable devices, such as Fitbit in epidemiological studies and the implementation of PA promotion policies in children.

However, this study has some limitations. First, because one of the study’s objectives was to examine compliance with wrist– and waist-worn accelerometers, the waist-worn accelerometer was used as the gold standard. Therefore, these findings should be compared with those obtained using more precise methods for assessing children’s activity; these include highly validated devices such as the ActiGraph (a wrist-worn accelerometer), ActivPAL (for measuring SB), and the combination of accelerometer data with daily activity logs [60]. Second, although a 1 min epoch length was used in this study, shorter (1–15 s) epoch lengths are recommended for children because the time spent on MVPA progressively decreases as the epoch length increases [61]. Therefore, this study’s results should be confirmed using shorter epoch lengths. Third, although our analysis was limited to measurement days on which the difference in wearing time between the two devices was less than 60 min, wearing time may considerably influence PA and SB estimates. Future studies should use wearable devices with heart rate monitors, as more detailed second-by-second wearing time can be confirmed by incorporating captured heart rate data [42]. Finally, the Fitbit Ace used in this study is the first model, with the Fitbit Ace 2 and 3 successors having already been released [62]. Because successor models may differ in terms of algorithms, comfort, and wearability, whether the results of this study apply equally to the successors and other wearable devices is unknown. Further research is needed to investigate the accuracy and precision of the Fitbit Ace PA evaluation estimates in free-living settings.

## Conclusions

Our results suggest that the Fitbit Ace is moderately correlated with waist-worn accelerometers for MPA, VPA, and MVPA measurements, and strongly to very strongly correlated for step count, SB, and LPA measurements. Furthermore, the Fitbit Ace exhibited significantly higher wear compliance than the waist-worn accelerometer. However, agreement was low across all indicators, and there were particular challenges in measuring VPA. Considering these results, although caution is required when using the Fitbit Ace for accurately measuring an individual’s PA, it appears to be useful tools for understanding the overall trends in children’s step counts, SB, and LPA patterns.

## Data Availability

All relevant data are within the manuscript.

## Acknowledgments

We are grateful to the participants who cooperated in the survey and to the school nurse teachers (yogo-kyoyu) and homeroom teachers in each school for conducting the survey. We are also truly grateful to the principal and vice principal of the elementary schools.

We would like to express our sincere gratitude to President and Representative Director Kazumichi Minato and Customer Associate Yusaku Watanabe from TechDoctor Inc. for their great efforts in acquiring the Fitbit data.

We would also like to express our sincere gratitude to Chief Executive Officer Narutoshi Tabuchi, Chief Knowledge Officer Megumi Tabuchi, Data Scientist Mitsuru Harada, and Yoji Yamamoto from AllegroSmart Inc. for their great support in acquiring the Fitbit data.

